# Long-term effects of extreme smoke exposure on COVID-19: A cohort study

**DOI:** 10.1101/2023.04.12.23288500

**Authors:** Tyler J. Lane, Matthew Carroll, Brigitte M. Borg, Tracy A. McCaffrey, Catherine L. Smith, Caroline Gao, David Brown, David Poland, Shantelle Allgood, Jillian Ikin, Michael J. Abramson

## Abstract

In early 2014, the Hazelwood coalmine fire covered the regional Australian town of Morwell in smoke and ash for 45 days. One of the fire’s by-products, PM_2.5_, has been linked higher rates of COVID-19 infection to increased expression of the ACE2 receptor, which the COVID-19 virus uses to infect cells throughout the body. However, it is unclear whether the effect persists for years after exposure. In this study, we surveyed a cohort established prior to the pandemic to determine whether PM_2.5_ from the coalmine fire increased long-term vulnerability to COVID-19 infection and severe disease.

In late 2022, 612 members of the Hazelwood Health Study’s adult cohort, established in 2016/17, participated in a follow-up survey including standardised items to capture COVID-19 infections, hospitalisations, and vaccinations. Associations were evaluated in crude and adjusted logistic regression models, applying statistical weighting for survey response and multiple imputation to account for missing data, with sensitivity analyses to test the robustness of results.

A total of 271 (44%) participants self-reported or met symptom criteria for at least one COVID-19 infection. All models found a positive association, with odds of infection increasing by between 4-21% for every standard deviation (12.3µg/m^3^) increase in mine fire-related PM_2.5_ exposure. However, this was not statistically significant in any model. There were insufficient hospitalisations to examine severity (*n*=7; 1%).

The findings were inconclusive in ruling out an effect of PM_2.5_ exposure from coalmine fire on long-term vulnerability to COVID-19 infection. Given the positive association that was robust to modelling variations as well as evidence for a causal mechanism, it would be prudent to treat PM_2.5_ from fire events as a risk factor for long-term COVID-19 vulnerability until more evidence accumulates.

## 1 Introduction

Ambient fine particles less than 2.5µm diameter (PM_2.5_) are associated with higher risk of COVID-19 infection, severe illness, and potentially death (1). One proposed mechanism is PM_2.5_ causing upregulation of Angiotensin-Converting Enzyme 2 (ACE2). The COVID-19 spike protein uses the ACE2 receptor to bind to and enter cells throughout the body (2,3). The combination of PM_2.5_ exposure and COVID-19 infection may exacerbate individual harms to cardiovascular and respiratory health (4,5).

Much of the existing evidence is drawn from studies of chronic ambient air pollution (1). There is limited direct evidence on whether extreme but discrete PM_2.5_ exposures like a coalmine fire increase long-term vulnerability to COVID-19. However, circumstantial evidence suggests this may be the case, such as higher expression of ACE2 in current *and* former smokers than non-smokers (6,7). And in Mexico City, PM_2.5_ measurements from 2000-2018 were associated with COVID-19 deaths in 2020, even when adjusting for PM_2.5_ exposures in 2019 and the weeks preceding infection (8).

Equivalent amounts of PM_2.5_ from landscape fires may be more hazardous than ambient urban background sources (9), meaning even short-term exposures could cause substantial long-term harm. Several ecological studies linked wildfire PM_2.5_ to increased COVID-19 infections (10–13), though these were essentially concurrent exposures and outcomes, separated by a few days or weeks. In New South Wales, Australia, areas with more burning during the 2019-2020 Black Summer bushfire season had higher population rates of COVID-19 infection in the following months, although there was no detectable association with coarser particulate matter air pollution (≤10µm, or PM_10_) (14).

In this study, we focused on the effects of the Hazelwood coalmine fire in regional south-eastern Australia, which covered the adjacent town of Morwell in smoke and ash for approximately six weeks in early 2014 (15). In the second half of 2022, we surveyed a cohort established after the coalmine fire (16) to address the following research question: Has PM_2.5_ exposure from the Hazelwood mine fire increased long-term risk of COVID-19 infection and severity?

## 2 Methods

### 2.1 Study population

The Hazelwood Health Study established an Adult Cohort to study the long-term effects of exposure to smoke from the Hazelwood coalmine. This consisted of 4,056 participants who were at least 18 years old during the coalmine fire and living either in Morwell (*n*=3,096), or nearby Sale (*n*=960), which had minimal exposure (16). In this follow-up, living participants for whom we had an email or phone number, had not participated in the concurrent psychological stream survey, and had not declined further contact were considered eligible, leaving 2,385 potential participants.

### 2.2 Data collection

Recruitment began in August 2022 and ended December 2022. Participants were invited to participate via either an email or text message, with several reminders. Study data were collected using REDCap (17,18), hosted by Monash University, and the Australian Eating Survey tool hosted at University of Newcastle (19).

### 2.3 Outcomes

A standardised questionnaire (20) was used to identify COVID-19 infections. For undiagnosed cases, participants were surveyed about a series of COVID-related symptoms in the years 2020 and 2021 and each month in 2022. Possible infections were determined using a standardised prediction formula based on symptoms and demographics (20,21).

Severe COVID-19 infections were indicated by self-reported hospitalisation.

### 2.4 Exposures

PM_2.5_ from the coalmine fire was retrospectively modelled with a meteorological and pollutant dispersion model with up to 100m^2^ resolution at hourly intervals (22). Individual exposures during the coalmine fire were estimated by combining PM_2.5_ concentration and time-location diaries from the original cohort survey (16).

### 2.5 Confounders

Demographic confounders included sex and age at survey. Health variables included Body Mass Index (BMI), calculated from current health and weight which were taken from the Australian Eating Survey, and having COPD. Vaccination was dichotomised as complete (4+) versus all other doses.

Socioeconomic factors were assessed with both area-based and individual-level indicators. The Index of Relative Social Advantage and Disadvantage (IRSAD) is a socioeconomic score for residential areas from the 2016 Australian census (23), which was linked to participant’s residence at Statistical Area level 2. Educational attainment was measured at the original cohort survey and grouped into “ secondary up to year 10,” “ secondary years 11-12,” and “ tertiary or trade qualification.”

As with socioeconomic status, we tested two indicators of tobacco use: smoker status and pack-years. Smoker status included “ current smoker,” “ former smokers” (≥100 lifetime cigarettes), and “ never smokers” (<100 lifetime cigarettes). Pack-years were the number of packs per day per year, where 1 pack year equalled 20 cigarettes per day for one year or 10 cigarettes a day for two years. Given the heavy positive-skew, pack-years were square root transformed.

### 2.6 Statistical analysis

Response/attrition bias was assessed using data measured at the initial Adult Cohort survey using the following variables: age at survey, sex, general health, low-SES residence (lowest quintile IRSAD score), residential area, educational attainment, and PM_2.5_ exposure.

Participant characteristics were summarised with descriptive statistics, grouped based on reported COVID-19 infection status. To account for differences between participating and non-participating cohort members, we applied inverse probability weights, balanced to represent the total sample of the follow-up survey. Missing data were addressed with multiple imputation by chained equations, generating 30 to correspond to the proportion of cases with missing data, and pooling results according to Rubin’s rules (24).

The effect of coalmine fire-related PM_2.5_ on COVID-19 infection/severity was tested using logistic regression models with several iterations: crude/adjusted, weighted/unweighted, and adjusting for location during the mine fire (exposure site Morwell or control site Sale). Due to concerns that PM_2.5_ effects on respiratory symptoms (25) would lead to false-positive COVID-19 cases on the symptoms rubric, we conducted sensitivity analyses on self-reported COVID-19 cases alone.

Coalmine fire-related PM_2.5_ exposure was standardised so that coefficients reflected a standard deviation change in particulate matter exposure. To ensure robustness of models, we tested several ways of adjusting for confounders. These included: 1) age as five-year groups or transformed into splines; 2) IRSAD as a continuous score or quintile-based groupings for the state of Victoria (low, middle three, and top as defined by the Australian Bureau of Statistics (23), which in our participants was effectively dichotomised because no participants resided in the top quintile of residential areas); and 3) smoking status or cigarette pack-years. Each confounder approach was tested in a fully-adjusted model, with the best-fitting selected based on Akaike Information Criterion (AIC).

Analyses were conducted in R (26) using RStudio (27) with the following packages: *gtsummary* (28), *haven* (29), *janitor* (30), *mice* (24), *naniar* (31), *readxl* (32), *see* (33), and *tidyverse* (34). Cleaning and analytical code are available on a public repository (35).

### 2.7 Ethics and funding

This study was approved as an amendment to the Hazelwood Adult Survey & Health Record Linkage Study (Project ID: 25680; previously CF15/872 – 2015000389 and 6066) by the Monash University Human Research Ethics Committee (MUHREC). Participants provided informed consent. This study was funded by the Department of Health, State Government of Victoria, though it represents the views of the authors and not the Department.

## 3 Results

### 3.1 Description of participants

Of 2385 invited cohort members, 612 (25.7%) completed the survey. Characteristics are summarised in Table 1. Almost half (44%) of respondents (*n*=271) either self-reported a COVID-19 infection (*n*=254, 42%) or met the symptom threshold (*n*=83, 14%). Only a quarter of self-report COVID-19 cases met the symptom threshold (*n*=66, 26%). Just seven respondents (1%) reported COVID-19-related hospitalisation.

**Table 1.** Descriptive statistics based on COVID-19 infection; continuous variables summarised with medians [inter-quartile ranges]

|  | COVID (*n* = 271) | No COVID (*n* = 341) | *p*-value |
| --- | --- | --- | --- |
| Hospitalised due to COVID | 7 (2.6%) | 0 (0%) | **0.003** |
| Mean fire-related PM<sub>2.5</sub> (µg/m<sup>3</sup>) | 8 [0-14] | 7 [0-14] | 0.541 |
| Resided in Morwell, during mine fire | 180 (67%) | 228 (67%) | 0.954 |
| Age at follow-up survey | 60 [50-68] | 66 [57-73] | **< 0.001** |
| Missing | 0 | 2 |  |
| Female | 165 (61%) | 197 (58%) | 0.345 |
| Smoking status |  |  | 0.221 |
| Non-smoker | 130 (49%) | 167 (49%) |  |
| Current smoker | 20 (7.5%) | 38 (11%) |  |
| Previous smoker | 118 (44%) | 133 (39%) |  |
| Missing | 2 | 4 |  |
| Cigarette pack-years | 0 [0-10] | 0 [0-20] | 0.156 |
| Educational attainment |  |  | **0.010** |
| Secondary up to year 10 | 49 (18%) | 78 (23%) |  |
| Secondary year 11-12 | 43 (16%) | 78 (23%) |  |
| Tertiary/trade+ | 177 (66%) | 184 (54%) |  |
| Missing | 1 | 2 |  |
| Socioeconomic status |  |  | 0.699 |
| Lowest residential quintile | 185 (70%) | 226 (68%) |  |
| Middle residential quintiles | 80 (30%) | 107 (32%) |  |
| Highest residential quintile | 0 (0%) | 0 (0%) |  |
| IRSAD residential score | 883 [812-932] | 865 [814-936] |  |
| Missing | 5 | 9 |  |
| Body Mass index (derived) | 29 [25-33] | 29 [26-34] | 0.440 |
| Height (metres) | 1.68 [1.62-1.75] | 1.68 [1.61-1.75] | 0.649 |
| Weight (kilograms) | 82 [70-95] | 84 [72-95] | 0.524 |
| Missing | 71 | 94 |  |
| Asthma/COPD | 97 (36%) | 133 (39%) | 0.415 |
| COVID-19 vaccination status |  |  | **< 0.011** |
| None | 9 (3.3%) | 20 (5.9%) |  |
| 1 | 3 (1.1%) | 1 (0.3%) |  |
| 2 | 36 (13%) | 44 (13%) |  |
| 3 | 139 (51%) | 90 (27%) |  |
| 4 | 83 (31%) | 183 (54%) |  |
| Missing | 1 | 3 |  |

There were little missing data aside from weight and height from the Australian Eating Survey, which were used to calculate BMI. As this was a separate questionnaire that required new registration and log-in, there was some loss to follow up, with only 73% (*n*=448) of participants completing it. In total, 30% (*n*=183) of participants had missing data, therefore we ran 30 imputations.

### 3.2 Response bias

Of 1846 non-participants, 1539 (79%) did not respond to the invitation, 238 (13%) refused, 67 (3.6%) had bounced emails and no phone number, 69 (3.7%) gave consent but did not complete the survey, and for 6 (0.3%) we were informed that the participant had died. The last group was likely an underestimate, since we were only informed that a cohort member had died when the invitation was sent to a shared account to which a living family member replied.

Cohort members were more likely to participate if they had better general health, higher educational attainment, and came from a higher socioeconomic area. While not significantly different, they were more likely to be female and have resided in Sale during the mine fire. Notably, there was no detectable difference in PM_2.5_ exposure. Comparisons between participating and non-participating cohort members are presented in Table S1. Inverse probability weights were calculated based on sex, age (modelled using natural splines), smoker status, general health at original survey, educational attainment, and the categorised residential socioeconomic indicator.

### 3.3 Association between PM_2.5_ and COVID-19 outcomes

Across 12 sets of models, all point estimates for the effect of PM_2.5_ from the coalmine fire on COVID-19 infections were positive (Odds Ratio range: 1.04-1.21 for a standard deviation of PM_2.5_, equivalent to 12.3µg/m^3^), although none were statistically significant. These are illustrated in Figure 1 and quantified in Table S2. With only 1% (*n* = 7) of participants reporting hospitalisation, regression analyses on this outcome were not feasible.

**Figure 1.**
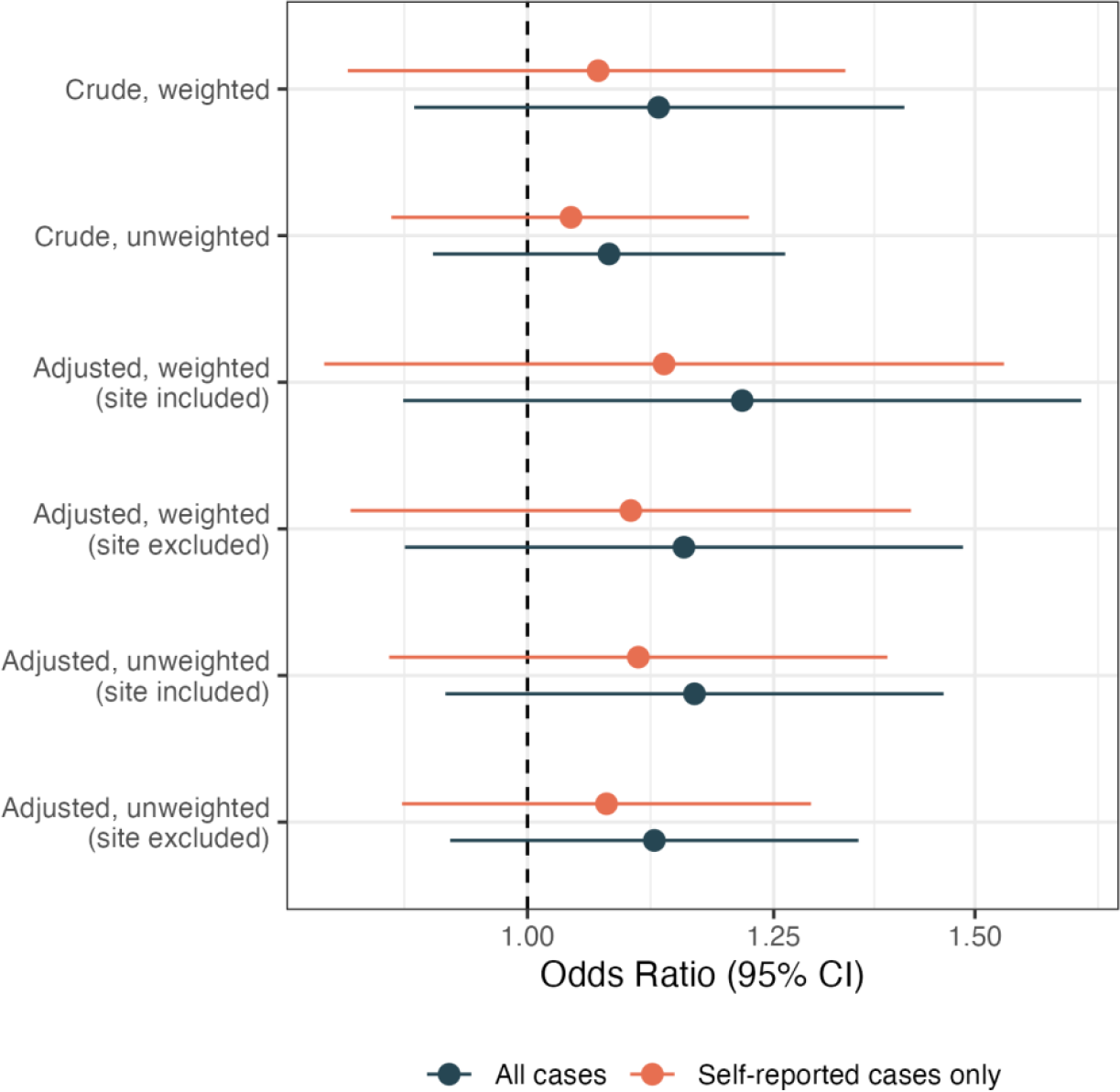
Association between standard deviation increase in PM_2.5_ exposure from the Hazelwood coalmine fire (12.3µg/m^3^) and COVID-19 infection under various modelling specifications

## 4 Discussion

PM_2.5_ exposure from the Hazelwood coalmine fire was positively but not significantly associated with COVID-19 infections. With due caution, we interpret this finding as inconclusive rather than evidence of no effect. There are several indicators that a real effect is plausible, including considerable mechanistic and epidemiological evidence that PM_2.5_ increases vulnerability to COVID-19 for a sustained amount of time (1–3,6,7). In addition, point estimates in our study were consistently positive across 12 separate models. The range of effects was smaller than observed in our systematic review, which found that a 10µg/m^3^ increase in PM_2.5_ was associated with a 66% increase in the odds of COVID-19 infection (1). Yet there are several reasons to expect a smaller effect in this instance. Studies in the systematic review focused on ambient PM_2.5_ only; none examined discrete exposures from events like coalmine or wildfires. While equivalent amounts of PM_2.5_ might be more harmful when originating from coalmine or wildfires (9), their discrete nature limits overall exposure.

Yet there remain reasons to doubt an association exists. The difference failed to meet the significance threshold of *p* ≤ 0.05, which, if anything, is too lax a standard (36). The gap between the coalmine fire and start of the pandemic was six years, a considerable amount of time for a medium- to long-term exposure to still impart an effect. Other findings from the Hazelwood Health Study have shown some recovery in respiratory function among adults measured in 2020 (PAPER FORTHCOMING). However, the proposed mechanism behind PM_2.5_ effects on poorer respiratory function is airflow obstruction (37), differs from the mechanism behind effect on COVID-19 vulnerability, which is an increase in ACE2 receptors. One may have a longer-lasting effect than the other.

There were too few hospitalisations to conduct a severity analysis in this sample. This is likely attributable to Australia’s response to the COVID-19 pandemic, especially in Victoria, which imposed stringent lockdowns until a high proportion of the population was vaccinated, limiting the number of severe COVID-19 cases.

### 4.1 Policy implications

Inconclusive results do not readily lend themselves to easy policy interpretations. Yet in the case of extreme PM_2.5_ and COVID-19 outcomes, it would be prudent to treat the association as real until determined otherwise. First, there is growing evidence that ambient PM_2.5_ increases risk of COVID-19 infection and severity (1), as well as concurrent smoke events such as wildfires (10–14). Even if discrete but extreme events like the Hazelwood coalmine fire do not increase longer-term vulnerability, it would still be highly beneficial to minimise shorter-term risks. Second, the list of health benefits of minimising PM_2.5_ exposure is large and growing (4); reducing the likelihood of longer-term COVID-19 risk only adds to them. Third, while the long-term health consequences of COVID-19 infections are not yet clear, it is likely that it leads to sustained complications throughout the body (38). Every averted COVID-19 infection, even within the same individual, can reduce the burden on the healthcare system, families, and society generally. In short, there is no harm to preventing extreme smoke events and otherwise limiting exposure, and a lot to be gained.

### 4.2 Strengths and limitations

This study has several limitations. The participation rate was modest and those who participated differed significantly from non-participants on several key factors. The COVID-19 symptom rubric used to identify undiagnosed cases was published in 2020 and, as indicated by the limited correspondence between self-report and symptom-identified cases, was probably outdated due to changes in the dominant variant. Symptoms in the rubric also overlapped with respiratory symptoms previously associated with PM_2.5_ exposure in this cohort (25), meaning smoke exposure could artificially inflate symptom-identified cases. However, there was no meaningful difference in results after restricting COVID-19 cases to those that were self-reported.

Among this study’s strengths are use of individual-level data from a cohort established several years before the COVID-19 pandemic, inverse probability weighting to account for response bias, and multiple imputation to account for missing data. Additionally, the study was pre-registered on the Open Science Framework (39), with research questions and a detailed statistical approach established prior to analysis and all deviations made explicit (see Appendix 1).

## 5 Conclusions

The findings were inconclusive as to whether PM_2.5_ from the Hazelwood coalmine increased long-term vulnerability to COVID-19. As climate change increases the frequency and intensity of wildfires, such as the kinds that led to the Hazelwood coalmine fire and the 2019-2020 “ Black Summer” that covered large parts of Australia in smoke, and the COVID-19 pandemic continues, there will be numerous opportunities to evaluate the long-term risk of extreme PM_2.5_ exposures. In the meantime, it would be prudent to treat long-term COVID-19 vulnerability as a real consequence of extreme smoke events and another reason to minimise exposure to PM_2.5_.

## Supporting information

STROBE checklist

## Data Availability

Data are confidential and cannot be shared publicly. However, we have archived cleaning and analytical code on a public repository.

https://doi.org/10.26180/22596991.v1

## 1 Appendix 1: Deviations from pre-registration

This study was pre-registered on the Open Science Framework on 19 July 2022 and updated during data collection on 8 November 2022 with a specific statistical analysis plan (1). There are several deviations from this plan, including: z-scoring PM2.5 exposure, square-root transformation of cigarette pack-years, dichotomising vaccination at 4+ doses, converting participant age with splines, IPW to account for non-response, and multiple imputation. Education groupings were not prespecified, but based on categories in previous Hazelwood Health Study research outputs (2). The asthma-COPD confounder variable was limited to just COPD as there is little evidence that asthma affects COVID-19 risk (3). Selection of final models based on Akaike Information Criterion was not pre-specified. Sensitivity analyses ignoring symptom-identified COVID-19 cases were added following early feedback raising concerns about false-positives due to symptoms from Hazelwood coalmine fire rather than COVID-19 infection. We initially planned to include PM2.5 data from the 2019-2020 “ Black Summer” wildfires”, though precise estimates were not available at the time of writing and preliminary analysis of air monitoring station data suggested little difference between Morwell and nearby areas.

## 2 Appendix 2: Black Summer bushfires

The original analysis plan called for inclusion of PM2.5 from the 2019/2020 Black Summer. However, the precise modelled data were not available at the time of writing. Further, air quality monitoring station data (4) suggested little difference between Morwell and the surrounding areas. This is illustrated in Figure S1, which shows daily mean PM2.5 estimates during the Black Summer in Morwell (combined from the east and south monitoring stations) and Rosedale, a town located between Morwell and Sale; Sale data were only available from 26 November to 12 December 2019, prior to the extreme exposures starting in January 2020. The time series suggest there was little meaningful difference between Morwell and surrounding areas.

## 3 Supplementary figures

**Figure S1.**
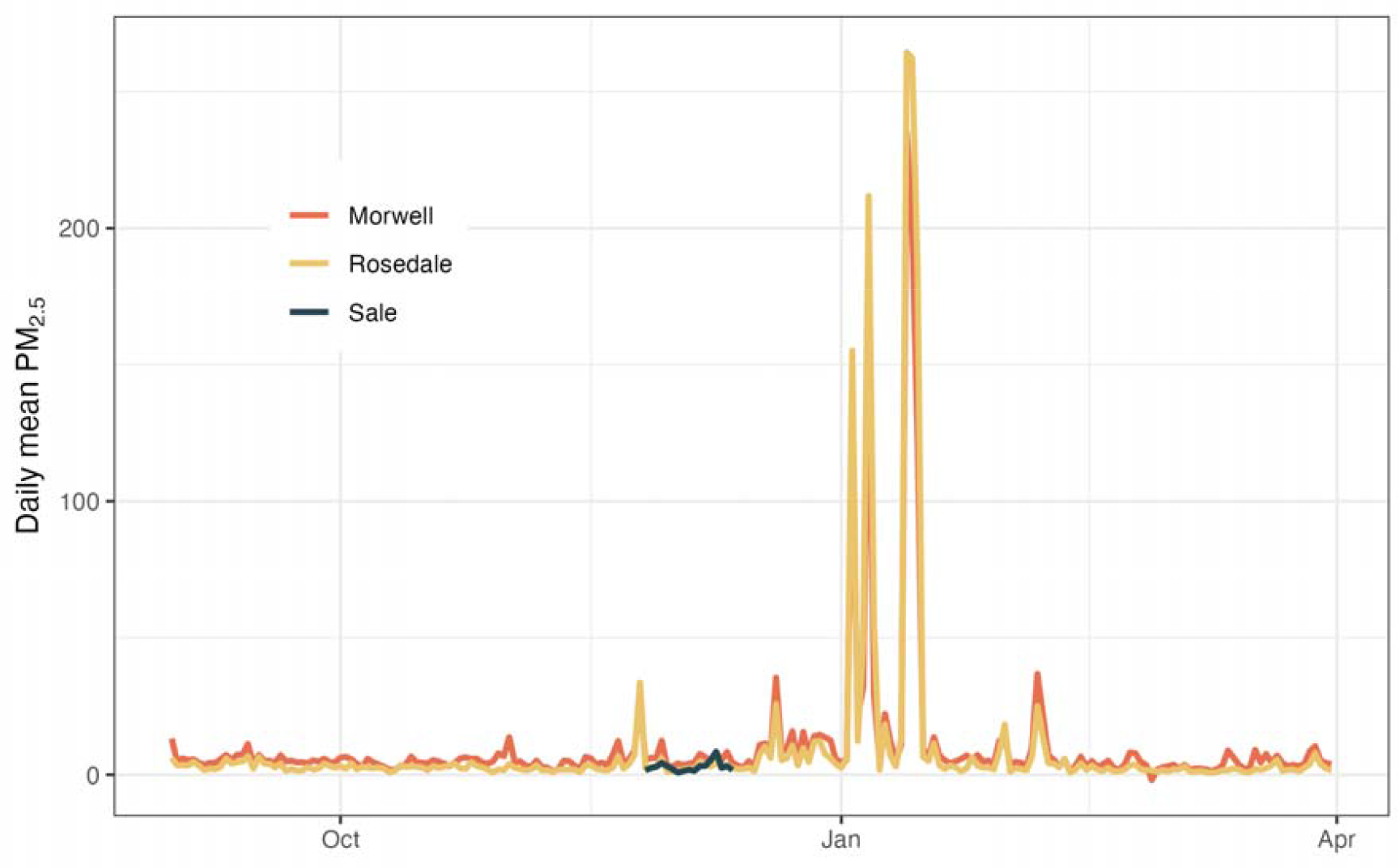
Comparison of daily mean PM_2.5_ in Morwell and Rosedale during the 2019/2020 Black Summer

## 4 Supplementary tables

**Table S1.** Differences between participating and non-participating cohort members

|  | Non-participating<br>(*n* = 1846) | Participating<br>(*n* = 612) | *p*-value |
| --- | --- | --- | --- |
| Female | 1000 (54.2%) | 362 (59.2%) | 0.056 |
| Age at original survey | 60 (IQR: 47-70) | 58 (IQR: 49-66) | **0.002** |
| Smoker status |  |  |  |
| Current smoker | **345 (19%)** | **74 (12%)** | **< 0.001** |
| Ex-smoker | **623 (34%)** | **249 (41%)** |  |
| Non-smoker | **873 (47%)** | **286 (47%)** |  |
| Missing | 5 | 3 |  |
| General health |  |  |  |
| Excellent/Very good | **705 (38.2%)** | **266 (43.5%)** | **0.004** |
| Good | **630 (34.1%)** | **220 (35.9%)** |  |
| Fair to poor | **497 (26.9%)** | **126 (20.6%)** |  |
| Missing | **14 (0.8%)** | **0 (0.0%)** |  |
| Educational attainment |  |  |  |
| Secondary up to year 10 | **584 (31.6%)** | **127 (20.8%)** | **< 0.001** |
| Secondary year 11-12 | **377 (20.4%)** | **121 (19.8%)** |  |
| Tertiary/trade+ | **868 (47.0%)** | **361 (59.0%)** |  |
| Missing | **17 (0.9%)** | **3 (0.5%)** |  |
| Residential socioeconomic area |  |  |  |
| Lowest quintile | 1378 (74.6%) | 411 (67.2%) | **< 0.001** |
| Middle three quintiles | 441 (23.9%) | 187 (30.6%) |  |
| Highest quintile | 0 (0.0%) | 0 (0.0%) |  |
| Missing | 27 (1.5%) | 14 (2.3%) |  |
| Residing in Morwell during the mine fire | 1301 (70.5%) | 408 (66.7%) | 0.076 |
| Mean fire-related PM<sub>2.5</sub> µg/m<sup>3</sup> | 8 (IQR: 0-14) | 8 (IQR: 0-14) | 0.338 |

**Note:** Dichotomous variables compared with Fisher's exact test, categorical with Pearson's Chi-squared test, and continuous variables with median (inter-quartile range), and Wilcoxon Rank Sum test

**Table S2.** Model results for effect of standard deviation (12.3µg/m^3^ ) increase in PM_2.5_ on likelihood of having at least one COVID-19 infection by the time of survey

| Model specifications |  |  | Self-reported and symptom-probable COVID-19 infections |  | Self-reported COVID-19 cases only |  |
| --- | --- | --- | --- | --- | --- | --- |
| Adjusted | Weighted | Site included | OR (95% CI) | *p*-value | OR (95% CI) | *p*-value |
|  | X |  | 1.13 (0.90-1.41) | 0.292 | 1.07 (0.85-1.33) | 0.575 |
|  |  |  | 1.08 (0.92-1.26) | 0.363 | 1.04 (0.88-1.22) | 0.632 |
| X | X |  | 1.15 (0.89-1.48) | 0.271 | 1.10 (0.85-1.42) | 0.469 |
| X |  |  | 1.12 (0.93-1.35) | 0.223 | 1.07 (0.89-1.29) | 0.448 |
| X | X | X | 1.21 (0.89-1.65) | 0.214 | 1.13 (0.83-1.54) | 0.43 |
| X |  | X | 1.16 (0.93-1.46) | 0.188 | 1.11 (0.88-1.39) | 0.383 |

## Notes

### Competing Interest Statement

The authors have declared no competing interest.

### Clinical Protocols

https://osf.io/yfgb3/

### Author Declarations

Monash University Human Research Ethics Committee approved this study as part of the Hazelwood Adult Survey & Health Record Linkage Study (Project ID: 25680; previously CF15/872 - 2015000389 and 6066)

